# Drive-through testing for SARS-CoV-2 in symptomatic health and social care workers and household members: an observational cohort study in Tayside, Scotland

**DOI:** 10.1101/2020.05.08.20078386

**Authors:** Benjamin Parcell, Kathryn Brechin, Sarah Allstaff, Meg Park, Wendy Third, Susan Bean, Chris Hind, Rajiv Farmer, James D Chalmers

## Abstract

It has been recognised that health and social care workers (HSCW) experience higher rates of infection with SARS-CoV-2. Widespread testing of HSCWs and their symptomatic household contacts (SHCs) has not been fully implemented in the United Kingdom. We describe the results of a testing programme for HSCWs and SHCs in a single UK region (Tayside, Scotland). The testing service was established 17 th March 2020 as the first in the country, and samples were collected at a drive-through testing hub based at a local community hospital. HSCWs with mild symptoms who were self-isolating and the SHCs of HSCWs who would therefore be absent from work attended for testing. From 17 th March 2020 to 11 th April, 1887 HSCWs and SHCs underwent testing. Clinical information was available for 1727 HSCWs and SHCs. 4/155 (2.6%) child contacts, 73/374 (19.5%) adult contacts and 325/1173 (27.7%) HSCWs tested positive for SARS-CoV-2. 15 of 188 undetermined cases were positive (8.0%). We estimate that testing prevented up to 3634 lost work days from HSCW testing, 2795 from adult SHC testing and 1402 lost work days from child SHC testing. The establishment of this testing programme has assisted the infection prevention and control team in their investigation of transmission and supported adequate staffing in health and social care sectors.

---

The novel coronavirus, designated SARS-CoV-2, has spread rapidly across the globe causing 1,850,220 confirmed cases at the time of writing and 114,215 deaths.^1^ Cases have been reported in 206 countries and COVID-19 has been declared as a pandemic by the World Health Organisation. The pandemic is causing substantial strain on healthcare systems worldwide due to a high frequency of affected cases requiring hospitalization and intensive care unit admission.^2^

Health and social care workers (HSCW) are disproportionately affected by infection with SARS-CoV-2. Healthcare workers accounted for 21% of cases of SARS during the 2002 outbreak^3^, and recent experience in China and Italy confirms high rates of healthcare worker infections during the current pandemic. In China, up until February 24^th^, 2055 healthcare workers were infected with 22 deaths.^4^ in Italy it has similarly been reported that 20% of documented cases are healthcare workers.^5^ In addition to the direct effects of the virus on HSCWs, infections have indirect effects on their families and staff morale and can have a significant impact on the ability of healthcare systems to function due to staff absence. Scotland recently began reporting staff absence rates in the health and social care sector showing that at the time of writing 1 in 20 staff were absent as a direct result of COVID-19.^6^

Current UK guidance for social distancing and self-isolation requires that HSCWs experiencing symptoms of a respiratory infection, such as cough or fever, should be absent from work for 7 days, while if a household contact is unwell, the staff member should be absent from work for 14 days to account for the incubation period of the virus. In most cases, particularly during the early stages of a pandemic, most respiratory illnesses will not be caused by SARS-CoV-2 and the rapid availability of testing would allow SARS-CoV-2 to be excluded and for critical healthcare staff to return to work. Nevertheless, widespread testing of HSCWs and their symptomatic household contacts (SHCs) has not been implemented in the United Kingdom or many other countries.

In this paper we describe the results of a testing programme for HSCWs and SHCs in a single UK region (Tayside, Scotland). The testing service was established 17^th^ March 2020 as the first in the country, and samples were collected at a drive-through testing hub based at a local community hospital. HSCWs with mild symptoms who were self-isolating and the SHCs of HSCWs who would therefore be absent from work were able to attend for testing. The testing model can be seen in Figure 1. Tayside Medical Science Centre (TASC) Research Governance are delegated responsibility for classification of audit, service improvement, service evaluation and healthcare research within Tayside. TASC confirmed that as per Health Research Authority (HRA) algorithm, this service evaluation did not require Sponsorship, Ethical review or NHS R&D Permission.

**Figure 1.**
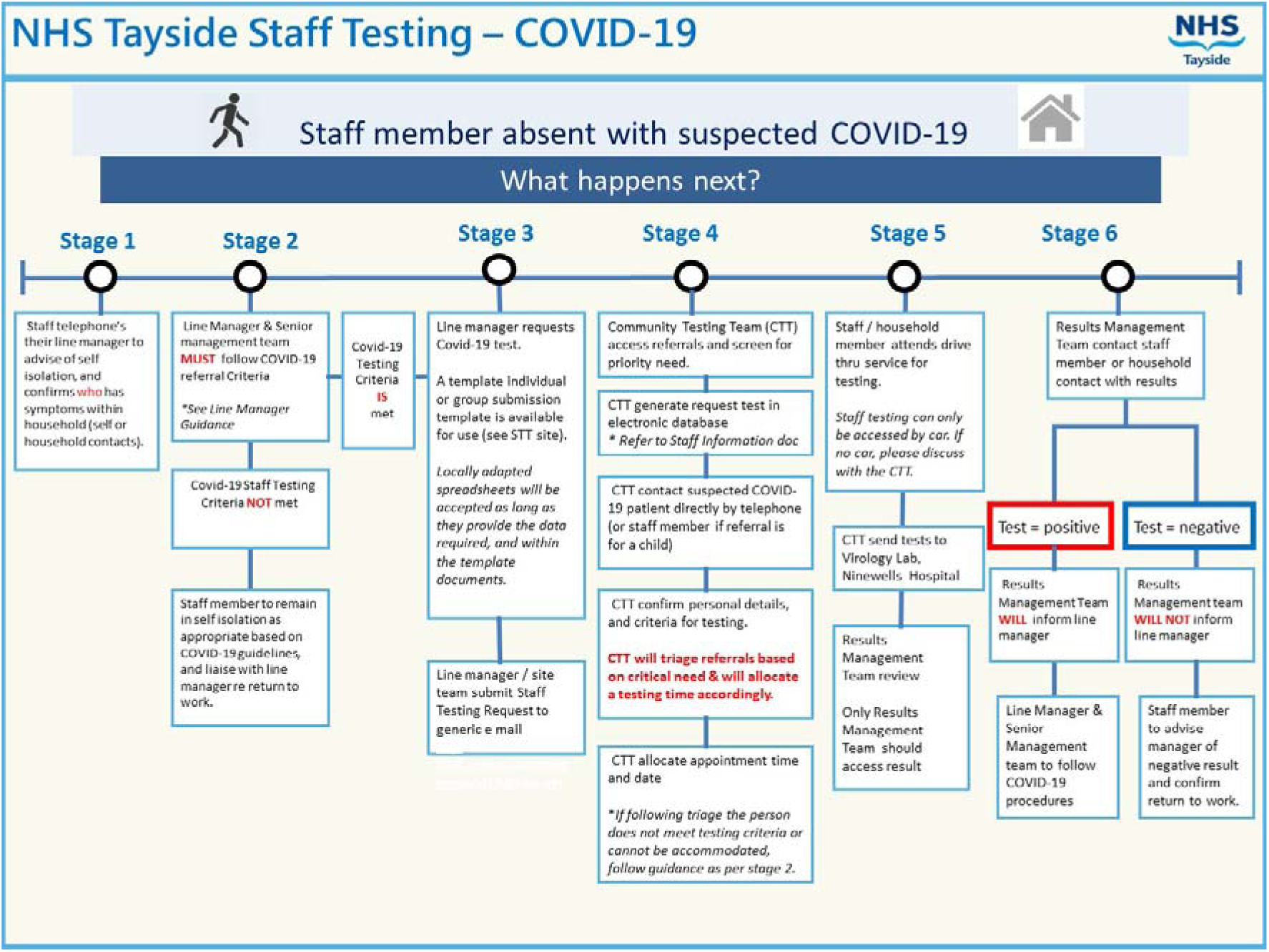
Process for requesting testing and reporting results.

Combined nasal and pharyngeal swabs were taken. In the laboratory nucleic acid was extracted and a one-step reverse transcription PCR assay (rtPCR) was carried out on an ABI7500 as previously described.^7^ HSCWs were able to return to work following a negative test with results available within 24 hours. Days of staff absence potentially saved were therefore calculated as the average number of working days following a negative test (up to a maximum of 6 days for staff members and 13 days in the case of SHCs).

From 17^th^ March 2020 to 11^th^ April, 1887 HSCWs and SHCs underwent testing. 3 individuals were tested more than once and therefore 1890 tests are included in the analysis. Clinical information was available for 1727 HSCWs and SHCs. The most frequently reported symptom was cough 1369/1727 (79.3%) followed by fever 619 (35.8%), sore throat 294 (17.0%) and shortness of breath 97 (5.6%). In 1173 cases (62.1%), the HSCW themselves was tested, in 374 cases (19.8%) an adult SHC was tested. In 155 cases a child SHC was tested (8.2%). In 188 cases (9.9%) it could not be determined whether a HSCW or SHC was tested.

4/155 (2.6%) child contacts, 73/374 (19.5%) adult contacts and 325/1173 (27.7%) HSCWs themselves tested positive for SARS-CoV-2. 15 of 188 undetermined cases were positive (8.0%). Based on the reported isolation time in each case and a typical 40 hour working week, we estimate that testing therefore prevented up to 3634 lost work days from HSCW testing, 2795 from adult SHC testing and 1402 lost work days from child SHC testing.

Assuming a worst case scenario in which all 188 undetermined cases were HSCWs, rather than SHCs, this equates to 8573 lost working days prevented through testing. All staff testing positive were isolated. At the time of writing there were no cases in which negative HSCWs subsequently tested positive for the disease or hospitalizations for suspected COVID-19 among HSCWs or SHCs tested in the community. No healthcare-associated outbreaks linked to staff that had been previously tested were reported during the study period.

To our knowledge, this is the first report of a systematic testing programme for HSCWs across a health board (Healthcare region) designed to ensure healthcare system resilience by reducing sickness absence. The results show a striking save of over 8000 lost working days for health and social care staff over a period of just 3 weeks which is likely to have a significant impact on the ability of health systems to respond to the SARS-CoV-2 pandemic. HSCW infections are a significant problem both from the point of view of delivering healthcare but also for patient safety as HSCWs may transmit the virus to vulnerable patients in hospital or care facilities. There have been few systematic studies of infections in healthcare staff, but one such study reported by Reuksen et al at two hospitals in the Netherlands found 4% of healthcare workers were infected.^8^ Most infections were mild and many staff continued to work due to the mild nature of the systems unaware they had SARSCoV-2.^8^ We speculate that availability and awareness of a staff testing programme for those with mild systems and their contacts may reduce the pressure for staff to work while unwell and therefore reduce the risks of healthcare-associated virus transmission. There are some important considerations when implementing a staff testing programme. Firstly, nasopharyngeal and oropharyngeal swabs have been reported to have a sensitivity ranging from 63% to 90%, but all studies suggest a sensitivity below 100%.^9,10^ It is therefore theoretically possible that some individuals tested in our study returned to work with a “falsenegative” result. We acknowledge this is a risk, but this small risk must be balanced against the risk of decimating the healthcare workforce through isolation of all HSCWs when they or their close contacts develop symptoms. We also acknowledge that we did not test for other respiratory viruses and that HSCWs or their household contacts may be infected with other communicable viruses, however household isolation for these reasons is not part of standard healthcare practice outside of pandemics. It is acknowledged that the precise number of working days saved through this practice is difficult to estimate as not all HCSWs would be working full time and on all days during the isolation periods which leads us to overestimate days saved. We have assumed a single HSCW per household but this is not necessarily correct and a SHC testing negative in some cases would result in more than one HSCWs being able to work, leading us to underestimate the days saved. Therefore the estimates provided should be treated with caution but likely represent the maximum utility of a healthcare working testing programme. The ability to perform over 1800 rtPCR tests for SARS-CoV-2 is only possible if there is sufficient testing capacity and it is acknowledged that this could be a limitation to implementing widespread testing in other regions or other countries. PCR testing for SARS-CoV-2 was only possible because of the availability of staff in the Virology laboratory in Tayside who are highly skilled in molecular diagnostic methods.

Nevertheless despite these limitations we report, to the authors knowledge, results of the first comprehensive regional testing programme for SARS-CoV-2. Maintaining adequate staffing of the social care sector, wards, intensive care units and primary care assessment units will become increasingly difficult as the pandemic progresses and we demonstrate that healthcare worker screening can markedly reduce the number of staff work days lost due to self-isolation.

## Data Availability

Data is stored by NHS Tayside

## Acknowledgements

The authors acknowledge the contribution of the staff at the NHS Tayside COVID-19 testing facility and the virology laboratory at Ninewells Hospital, Dundee.

## References

1 European Centres for Disease Control, https://www.ecdc.europa.eu/en/publicationsdata/download-todays-data-geographic-distribution-covid-19-cases-worldwide accessed 13th April 2020

2 Guan W-J, Ni Z-Y, Hu Y, et al. Clinical Characteristics of Coronavirus Disease 2019 in China. N Engl J Med. February 2020:10.1056/NEJMoa2002032. doi:10.1056/NEJMoa2002032

3 World Health Organization (2003) Case definitions for surveillance of severe acute respiratory syndrome (SARS). WHO, Geneva. (https://www.who.int/csr/sars/casedefinition/en/) accessed 13th April 2020

4 World Health Organisation. Report of the WHO-China Joint Mission on Coronavirus disease 2019 (COVID-19).; 2020. https://www.who.int/docs/defaultsource/coronaviruse/who-china-joint-mission-on-covid-19-final-report.pdf. Assessed 13^th^ April 2020.

5 Remuzzi A, Remuzzi G. COVID-19 and Italy: What Next. Lancet 2020; 395(10231):1225–1228

6 https://www.gov.scot/coronavirus-covid-19/ accessed 13th April 2020

7 https://www.england.nhs.uk/coronavirus/wpcontent/uploads/sites/52/2020/03/guidance-and-sop-covid-19-virus-testing-in-nhslaboratories-v1.pdf

8 Reuksen CB, Buiting A, Bleeker-Robers C et al. Rapid Assessment of Regional SARS-CoV-2 Community Transmission Through a Convenience Sample of Healthcare Wokers, The Netherlands, March 2020. Euro Surveill 2020;25(12).

9 Loeffelholz MJ, Tang YW. Laboratory diagnosis of emerging human coronavirus infections-the state of the art. Emerg Microbes Infect 2020;9(1):747–756

10 Wang W, Xu Y, Gao R et al. Detection of SARS-CoV-2 in different types of clinical specimens. JAMA 2020; Mar 11 online ahead of print.

